# Transcranial Magnetic Stimulation Measures of Corticospinal Excitability in Black and Hispanic/Latino People with Painful Peripheral Neuropathy

**DOI:** 10.1101/2025.01.08.25320215

**Authors:** Marlon L. Wong, Lisa McTeague, Gabriel Gonzalez, Juan Gonzalez, Jessica Bolanos, Peter J. Hosein, Danylo F. Cabral, Peter J. Fried

## Abstract

This study aims to provide preliminary descriptive data on transcranial magnetic stimulation (TMS) measures obtained in Black and Hispanic/Latino individuals with chronic painful peripheral neuropathy (PN), including those with chemotherapy-induced peripheral neuropathy (CIPN) and diabetic neuropathy (DN). CIPN and DN share similar neuropathic symptoms and underlying physiological mechanisms, in particular altered central nervous system processing. TMS is a non-invasive technique that can assess corticospinal excitability and the function of GABAergic and glutamatergic pathways, potentially serving as a diagnostic tool for PN. This secondary analysis utilized data from a pilot randomized sham-controlled trial of transcutaneous auricular vagus nerve stimulation (taVNS) in people with PN. TMS measures, including resting motor threshold (RMT), unconditioned MEP amplitude (spTMS), short interval intracortical inhibition (SICI), and intracortical facilitation (ICF), were assessed at baseline over 2 separate visits. Test-retest reliability was evaluated, and changes in TMS measures following transcutaneous auricular vagus nerve stimulation were analyzed. Baseline TMS measures showed no significant differences between CIPN and DN groups. The study found good test-retest reliability for TMS measures, with ICC values between 0.73 and 0.87 for all TMS measures. Post taVNS, neuropathic pain symptoms improved, with a slight decrease in ICF. Overall, TMS measures demonstrated good reliability in this sample of Black and Hispanic/Latino individuals with PN, and these findings provide valuable preliminary data for future studies aimed at establishing the psychometric properties and diagnostic utility of TMS measures in PN.

## INTRODUCTION

Peripheral neuropathy (PN) is the most common neurodegenerative disorder, and its prevalence is increasing due to two major causes, the increase in survivorship among cancer patients treated with neurotoxic chemotherapies and the expanding epidemic of diabetes (1, 2). It is estimated that up to 90% of people who receive neurotoxic chemotherapies will develop acute chemotherapy-induced peripheral neuropathy (CIPN) and over 30% develop chronic CIPN (3, 4). Similarly, PN is believed to affect up to 50% of people with diabetes (DN), causing pain and reducing quality of life (5, 6). PN is also known to place greater burden on racial and ethnic minority communities in the United States, particilarly for Black, Hispanic/Latino, and Native American communities (7, 8).

Although the precipitating factors that lead to CIPN and DN differ, both are characterized by distal symmetric dysesthesias and paresthesias in glove/stocking distributions. Further, the associated physiological mechanisms believed to underly the perpetuation of neuropathic symptoms are similar for both groups (i.e., neuroinflammation, autonomic dysregulation, and altered central nervous system processing) (9–11). A review of both human and animal studies suggested that CIPN is partly caused by brain hyperactivity and reduced GABAergic inhibition (12). Specifically, neurotoxic chemotherapies have been shown to cause a decrease in GABA in the thalamus (13), and restoration of GABA levels, or experimentally activating the GABA pathway, reduces CIPN symptoms (14–18). Likewise, DN is known to be associated with reduced GABAergic inhibition and increased thalamic excitability (19–22).

Therefore, non-invasive measurement of GABA in the central nervous system might serve as a mechanistic endpoint for clinical trials for PN, and they could direct the development of non-invasive interventions to maximize GABAergic signaling through endogenous mechanisms (23). Transcranial magnetic stimulation (TMS) is a promising solution as it can be used to assess corticospinal excitability via the function of GABAergic and glutamatergic and serotonergic pathways in the motor cortex. The clinical diagnostic utility of TMS techniques have been reported across a range of diseases, including neurodegenerative, inflammatory, and lesional brain and spinal disorders (23). Although studies have demonstrated reduced corticospinal-motor plasticity in people with diabetes using TMS (24–26), these techniques have not been applied to characterize neurophysiological changes associated with CIPN or DN. Prior studies on the role of the central nervous system in PN have largely focused on imaging technologies, with only a few using electroencephalogram (EEG) or electromyography (EMG) for H-reflex assessement, and to our knowledge none have explored the use of TMS measures as diagnostic tools (12, 19).

Emerging evidence suggests that PN symptoms are accompanied by both structural and functional changes in the brain, namely in the pain modulation areas (27). Additionally, both CIPN and DN have been shown to be partly caused by corticospinal hyperactivity and reduced GABAergic inhibition (12, 14–22). Thus, non-invasive brain stimulation techniques, such as transcutaneous auricular vagus nerve stimulation (taVNS), are promising tools for managing PN and warrant exploration. taVNS is particularly intriguing because it is known to act on multiple mechanisms that contribute to PN, such as dysregulation of the autonomic nervous system (28–30), inflammation (31), and structural and functional changes in the brain (27). TMS measures may be a used to probe resulting neurophysiological changes with taVNS and thus improve understanding of the underlying mechanisms for the effects of taVNS on PN. Before this can be done, the feasibility of conducting these measures, and the psychometric properties and normative data on these measures, must be established in this population. Further, Black and Hispanic/Latino people have historically been severely underrepresented in TMS research (32), despite experiencing a higher prevalence of CIPN and DN (7, 8). It is important to include these communities in this research to ensure that the research reaches all of those who may benefit and to make research findings more generalizable. Thus, the purpose of this study was to provide preliminary and detailed descriptive data on TMS measures in a cohort of Black and Hispanic/Latino patients with chronic PN.

## METHODS

This study is a secondary analysis of data from a pilot randomized sham-controlled trial (RCT) that was designed to examine the influence of educational videos on transcutaneous auricular vagus nerve stimulation (taVNS) on participant expectations for pain relief with taVNS in Black and Hispanic/Latino people with CIPN (n=17) or DN (n=11). A detailed description of the parent study is provided elsewhere (clinicaltrials.gov #NCT05896202) (33). Briefly, participants were randomly assigned to video (intervention) or control groups, and all participants completed 3 visits (**Figure 1**): The first visit consisted of ∼90 minutes of education on both taVNS and TMS, including review of brochures and consent forms (both groups) and 3 short video segments on taVNS for the intervention group; the second visit consisted of a baseline assessment battery, which included TMS measures, followed by a 60-minute active taVNS session for the intervention group and randomly assigned active or sham taVNS for the control group, and then the assessment battery was repeated; the 3^rd^ visit consisted of only the assessment battery for the intervention group, and the control group were crossed over such that those who received the active taVNS on visit 2 received sham on visit 3, and the assessment battery was administered pre and post active or sham taVNS. We did not anticipate that a single session of taVNS would have lasting effects, and participants were required to schedule visit 3 at least 48 hours after visits 2. Thus, baseline TMS measures at visits 2 and 3 were used to assess test-retest reliability of TMS measures.

**Figure 1.**
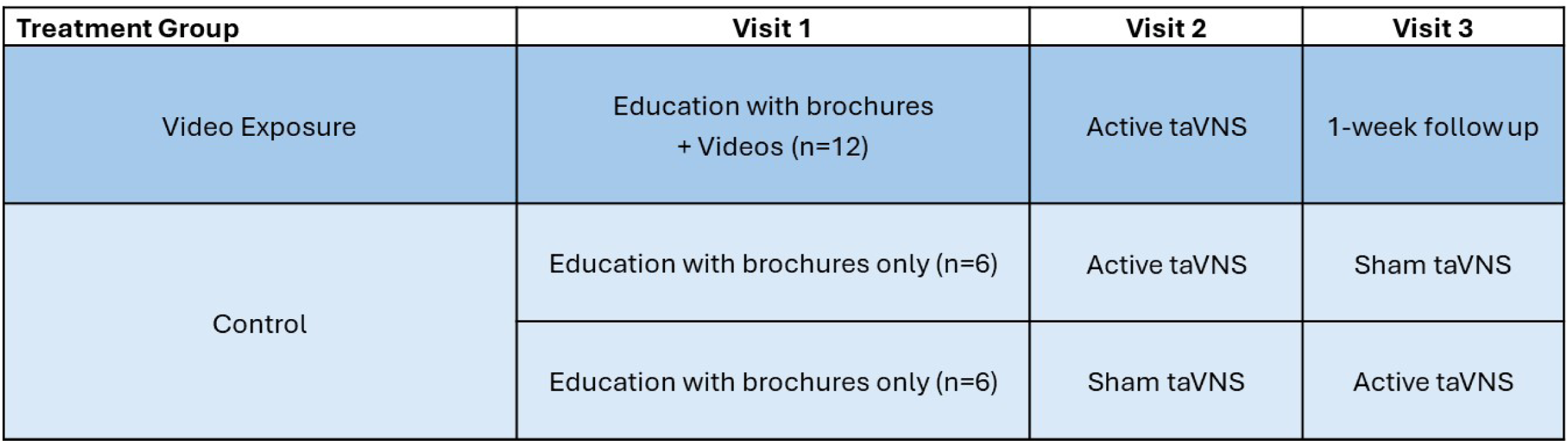
Pilot Sham-Controlled RCT Study Design

### Participants

Participants were recruited from the University of Miami medical health care system from January to May 2024. Potential participants were identified by medical record and then their respective providers (i.e., oncologist or endocrinologist) informed them about the pilot study during clinical visits. Inclusion criteria included anyone with glove or stocking distribution paresthesia or dysesthesia that developed after receiving neurotoxic chemotherapies, or with a medical diagnosis of DN, and who self-identified as Black or Hispanic/Latino and had a score on the Neuropathic Pain Symptom Inventory(34) greater than 10. Exclusion criteria included 1) any unstable medical condition or medical contraindication to moderate physical exertion (e.g., unstable angina or cardiac arrythmia), 2) pregnancy, 3) presence of cognitive impairment or language barrier that impairs full autonomy in the consent process or in the ability to participate in detailed interviews, 4) implants in the head or neck, cochlear implants, or pacemaker, 5) head or neck metastasis or recent ear trauma, 6) history of seizures. Approval was granted by the University of Miami Institutional Review Board, and informed consent was obtained from all participants.

### Procedures

In the first visit, all participants were educated on taVNS and TMS using brochures and the consent form, and the intervention group received video enhanced education on taVNS. The education included discussion on the use of TMS as an intervention as well as an assessment tool for research purposes, the proposed mechanisms of action for taVNS and TMS, and the risks and regulations associated with taVNS and TMS research. Racial and ethnic differences between participant and investigators/providers is also known to influence expectations and pain outcomes (35, 36); thus, a Black investigator provided all educational sessions for Black participants, and a Hispanic/Latino investigator provided all education sessions for Hispanic/Latino participants and in their preferred language (English or Spanish). All participants had ample opportunity to ask questions and discuss the content with the investigators.

At the end of the educational session, participants provided feedback on the educational materials and completed the EXPECT(37) questionnaire. The EXPECT is a 4-item questionnaire that assesses expectations for pain improvement. Each of the 4 items is scored on an 11-point scale, with 0 being no change and 10 representing complete relief (37). The mean of the 4 items is presented as the EXPECT score. For the second and third visits, all participants received the same TMS measures at baseline and post active or sham taVNS.

### TMS resting motor threshold (RMT), unconditioned single pulse TMS (spTMS), short-interval intracortical inhibition (SICI), and intracortical facilitation (ICF)

TMS has been used in neuroscience research for over 40 years, and a wide variety of diagnostic approaches have been developed (23). However, there are 4 TMS diagnostic approaches that we believe are of particular interest to PN researchers and clinicians given their relative simplicity and potential value:

#### 1. Motor Threshold

When TMS is applied to the motor cortex (M1) with sufficient stimulation intensity, motor evoked potentials (MEPs) can be recorded from contralateral extremity muscles (38). The lowest intensity required to consistently evoke MEPs in the target muscle is referred to as the motor threshold (MT). While MT is known to be an indicator of short-lasting glutamatergic AMPA transmission in corticospinal neurons (23, 39, 40), it is important to note that MT is also result of the combined excitability of 1) the core of neurons that represent the target muscle in M1, 2) the interneurons projecting onto these neurons, 3) motor neurons in the brainstem and spinal cord, and 4) neuromuscular junctions and muscle (23, 38, 40). Thus, MT provides insights on the efficacy of the entire pathway being investigated (e.g., from the presynaptic cortical neurons to the target muscle) (38).

#### 2. MEPs with unconditioned single pulse TMS (spTMS)

Cortical excitability is also represented by the amplitude of MEPs elicited by a single suprathreshold (delivered at an intensity above MT) TMS pulse stimulus. However, there is great inter and intra-individual variability with single pulse MEP amplitudes, and therefore this measure is often considered qualitative rather than quantitative (38).

#### 3. Short latency intracortical inhibition (SICI)

SICI involves stimulating the primary motor cortex (M1) with a subthreshold conditioning-stimulus followed by suprathreshold test-stimulus at inter-stimulus intervals (ISI) of 1-to-6 ms. This typically results in a decreased MEP amplitude compared to the test-stimulus alone (23), and pharmacological studies have suggested SICI is mediated by inhibitory inter-neuronal circuits acting via GABA-A receptors (41, 42).

#### 4. Intracortical facilitation (ICF)

Using the same parameters as SICI while simply increasing the ISI to 8-30 ms results in the opposite effect: an increased MEP amplitude compared to the test-stimulus alone (23). This technique is termed intracortical facilitation (ICF), and it is an NMDA receptor dependent phenomenon that is modulated by serotonergic neurotransmission (43, 44). Thus, both SICI and ICF examine corticospinal excitability but through different mechanisms.

For this study, TMS measures were assessed using the Magpro X100 (MagVenture, Alpharetta, GA). Participants were seated in an armchair with the head on a headrest while TMS was delivered through a focal figure-of-eight shaped magnetic coil (MC-B70, MagVenture), and motor evoked potentials (MEP) were recorded using surface EMG Ag/AgCl electrodes placed over the right abductor digiti minimi muscle in a belly-tendon montage. The coil position leading to the highest peak-to-peak amplitude of the MEP (‘hotspot’) was marked using the TMS navigation system (Localite, Germany) to ensure accurate coil positioning throughout the testing. A fixed sequence of TMS measurements were followed:

1. First, resting motor threshold (RMT) was defined as the lowest stimulator output intensity that induced MEP peak-to-peak amplitude greater than 50 μV in five out of 10 consecutive trials.
2. Then the single pulse (at 130% of RMT) and paired-pulse parameters, SICI and ICF, were obtained in a pseudo-random order. For paired pulse, the conditioning-stimulus was set to an intensity of 70% of the RMT, and the test-stimulus was set to 130% of RMT. We covered a range of interstimulus intervals (ISIs) between SICI and ICF: including 1, 2, and 3 milliseconds (ms) for SICI, and 7, 10, and 15ms for ICF, with 10 pulses at each interval. By covering a range of ISIs our intent was to minimize the risk of missing the peak ISI for a given individual. The average of the 10 trials was used to define the amplitude of the peak-to-peak MEP for each condition, and the mean of all trials at ISIs 1, 2, and 3 (30 pulses) was used for SICI, and the mean of all trials at ISIs 7, 10, and 15 (30 pulses) was used for ICF.

RMT is presented as a percentage of machine maximum output, and spTMS is presented in millivolts (mV). SICI and ICF values are presented as the relative percentage of mean paired pulse MEP values to mean spTMS MEP values, minus 100 [(conditioned response/unconditioned response) × 100%)-100], such that negative values represent a relative inhibitory effect, and positive values represent a relative faciliatory effect.

### Neuropathic Pain Assessment

The Neuropathic Pain Symptom Inventory (NPSI)(34) was administered as part of the screening process. The NPSI is one of the most widely used tools for characterizing neuropathic pain symptom severity (45, 46), and it assess dimensions of neuropathic pain (burning spontaneous pain, pressing spontaneous pain, paroxysmal pain, evoked pain, and paresthesia/dysesthesia) over the last week. NPSI total score ranges from 0 to 100, with higher scores indicating worse NP severity. Additionally, at each visit participants were asked to rate the severity of their symptoms at the immediate moment with 0-10 numeric rating scales for pain, numbness, tingling, burning, and shooting/electric shocks; this was asked both before and after taVNS stimulation.

### Analyses

Descriptive statistics (i.e., means, medians, and standard deviations) were calculated for all TMS measures. Group comparisons between CIPN and DN groups were made using Mann-Whitney U or Chi-Square tests, and changes pre post taVNS were assessed with Wilcoxon Signed Ranks tests, with α set at 0.05. Intraclass Correlation Coefficient (ICC) analysis, using an alpha two-way mixed model for absolute agreement, was used to assess the reliability of TMS measures and reported with 95% confidence intervals. All statistical analyses were conducted using Statistical Package for the Social Sciences (SPSS) v28 (IBM Corp., Armonk, NY) and figures were rendered using GraphPad Prism v9.3.1 (GraphPad Software, La Jolla, CA).

## RESULTS

Twenty-eight participants were enrolled (17 with CIPN and 11 with DN); however, only 24 completed both visits 2 and 3, with 23 included in partial data analysis and 19 with complete TMS data (**Figure 2**-CONSORT). Five participants were missing TMS data because their RMT exceeded 77% of the machine’s maximum output, and therefore a stimulus intensity of 130% of RMT could not be achieved for single or paired pulse assessments. Demographic and visit 1 pain information for the participants can be found in **Table 1**. Participants with DN had slightly higher symptoms on average than the participants with CIPN, but there were wide ranging scores across both groups and no meaningful differences between them (all p values >0.14). Baseline scores at visit 2 and 3 can be found in **Figure 3**.

**Figure 2.**
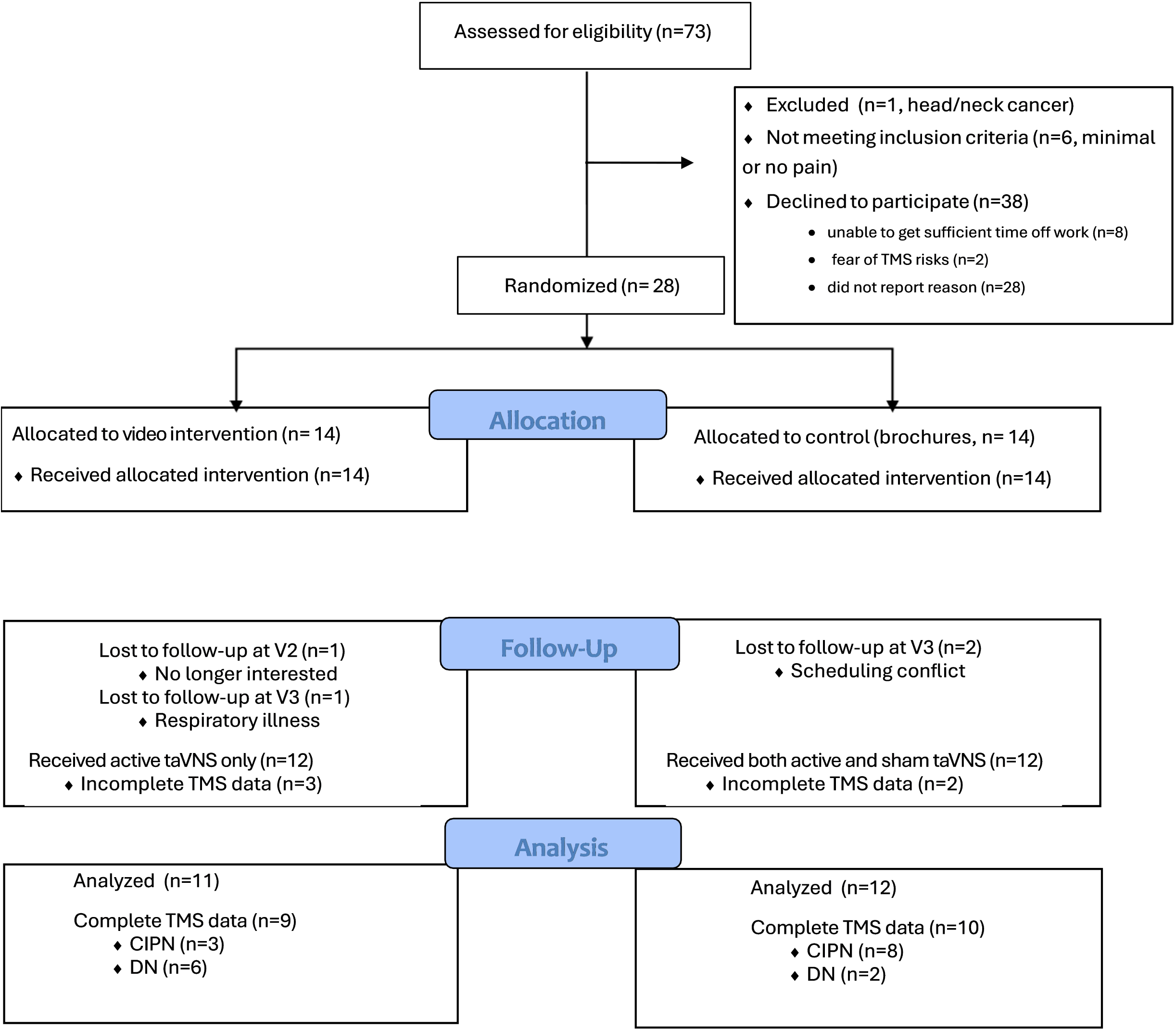
CONSORT Diagram

**Table 1.** Demographic and Pain Characteristics

|  | ***CIPN***<br>***(n=14)*** | ***DN***<br>***(n=9)*** | ***Total***<br>***Sample***<br>***(n=23)*** | ***P value*** |
| --- | --- | --- | --- | --- |
| *Age\** | 60.4 (9.2) | 54.4 (8.2) | 58.0 (9.1) | 0.179 |
| *Gender (%Female)* | 78.6 | 66.7 | 73.9 | 0.526 |
| *Race (% Black)* | 42.9 | 77.8 | 56.5 | 0.058 |
| *Ethnicity (% Hispanic)* | 71.4 | 66.7 | 69.6 | 0.809 |
| *Using pain medication (%Yes)* | 85.7 | 55.6 | 73.9 | 0.108 |
| • *Duloxetine (n)* | 0 | 1 | 1 | 0.202 |
| • *Gabapentin (n)* | 10 | 5 | 15 | 0.435 |
| • *Opioids (n)* | 0 | 1 | 1 | 0.202 |
| *Pain\* (0-10)* | 6.9 (2.5) | 8.3 (1.1) | 7.5 (2.2) | 0.130 |
| *Numbness\* (0-10)* | 7.8 (1.9) | 8.6 (1.3) | 8.1 (1.7) | 0.351 |
| *Tingling\* (0-10)* | 7.8 (2.0) | 8.0 (1.6) | 7.9 (1.8) | 0.898 |
| *Burning\* (0-10)* | 6.6 (3.2) | 5.6 (3.9) | 6.2 (3.4) | 0.567 |
| *Shooting\* (0-10)* | 5.9 (3.3) | 7.7 (3.3) | 6.6 (3.3) | 0.127 |
| *NPSI Score\* (0-100)* | 57.7 (20.1) | 68.7 (19.2) | 62.0 (20.1) | 0.122 |
| *EXPECT Score\* (0-10)* | 8.0 (2.0) | 7.5 (1.2) | 7.8 (1.7) | 0.207 |

*\*Mean (standard deviation)*

**Figure 3.**
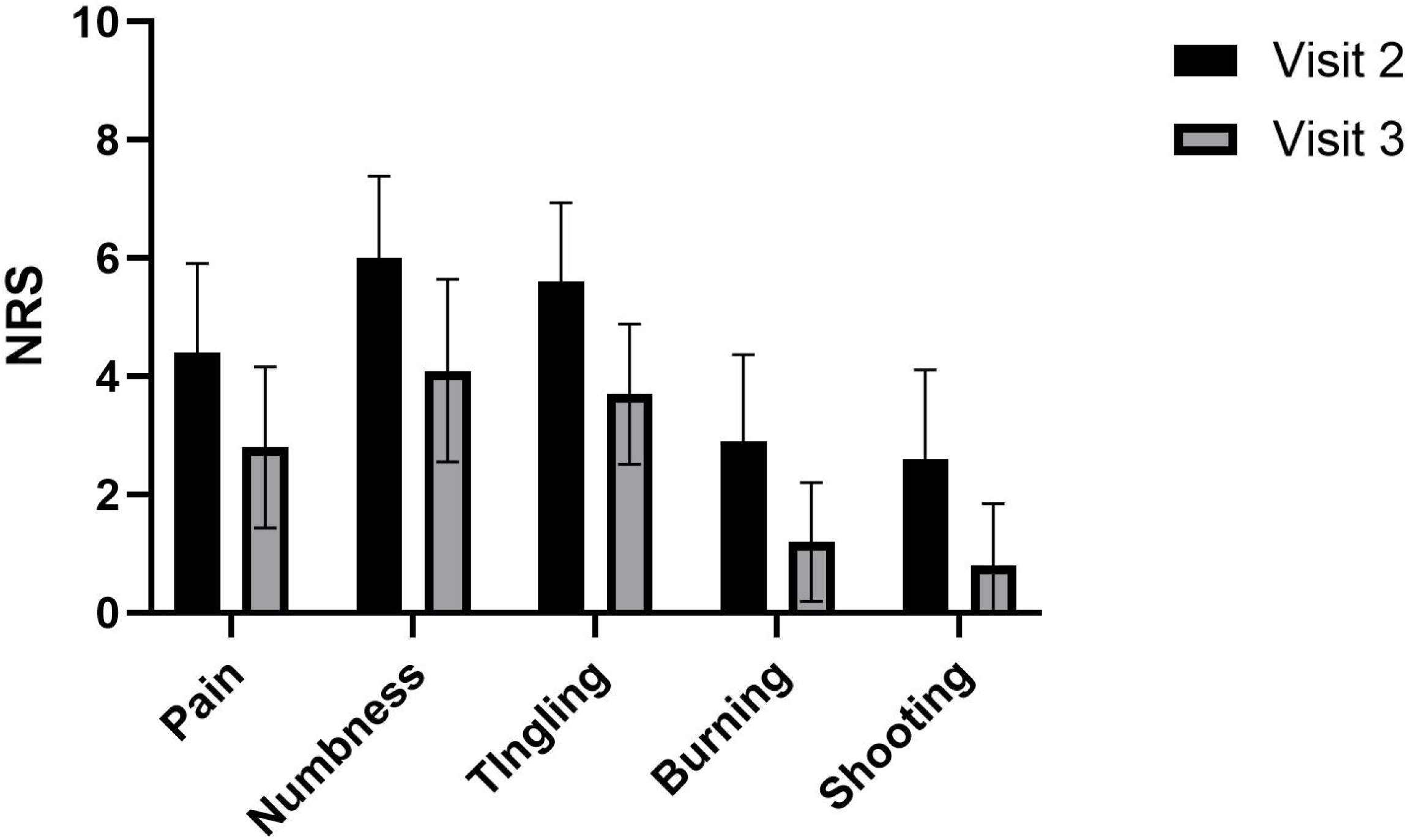
Baseline Symptoms at Visits 2 and 3 *Mean with 95% Confidence Interval; NRS=numeric rating scale*

### Baseline TMS Measure Descriptives

There were no significant differences in baseline TMS measures at visit 2 between participants with CIPN and those with DN, with median values for the entire sample of 70.0 for RMT, 0.87mV for SP, -40.9% for SICI, and 10.1% for ICF (**Table 2**). Additionally, there were no significant differences in baseline values between visits 2 and 3, with mean differences of -0.05 for RMT, 0.01mV for SP, 4% for SICI, and -9% for ICF.

**Table 2.**
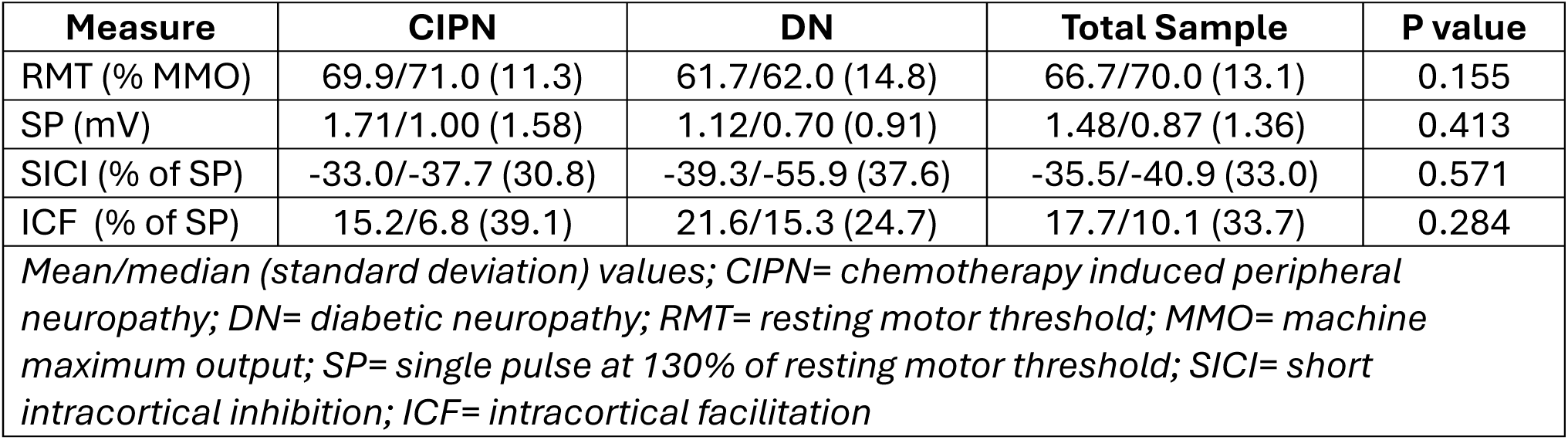
Visit 2 TMS Measures at Baseline

| Measure | CIPN | DN | Total Sample | P value |
| --- | --- | --- | --- | --- |
| RMT (% MMO) | 69.9/71.0 (11.3) | 61.7/62.0 (14.8) | 66.7/70.0 (13.1) | 0.155 |
| SP (mV) | 1.71/1.00 (1.58) | 1.12/0.70 (0.91) | 1.48/0.87 (1.36) | 0.413 |
| SICI (% of SP) | -33.0/-37.7 (30.8) | -39.3/-55.9 (37.6) | -35.5/-40.9 (33.0) | 0.571 |
| ICF (% of SP) | 15.2/6.8 (39.1) | 21.6/15.3 (24.7) | 17.7/10.1 (33.7) | 0.284 |
| *Mean/median (standard deviation) values; CIPN= chemotherapy induced peripheral neuropathy; DN= diabetic neuropathy; RMT= resting motor threshold; MMO= machine maximum output; SP= single pulse at 130% of resting motor threshold; SICI= short intracortical inhibition; ICF= intracortical facilitation* |  |  |  |  |

### Test-retest reliability of TMS measures

The median number of days between visits 2 and 3 was 5 (with a mean of 6.2 and a range of 2-26 days). Mean NRS scores for all symptoms decreased slightly from visit 2 to visit 3 (mean change scores ranged from -1.6 to -0.8), and there was no difference between the participants with CIPN and those with DN (p values > 0.10). For the CIPN group, there was fair test-retest reliability for ICF (ICC=0.67, CI=-0.34 to 0.91, p=0.057) and good test-retest reliability for SICI (ICC=0.84, CI=0.40 to 0.96, p=0.005). ICC values for individual paired pulse intervals ranged from 0.63 to 0.84 (**Figure 4A**), and RMT and SP had ICCs of 0.6 (CI=-0.52 to 0.89, p=0.083), and 0.79 (CI=0.28 to 0.94), respectively. The DN group demonstrated excellent reliability for all TMS measures with ICC values ≥0.93 (**Figure 4B** and **Table 3**).

**Figure 4A.**
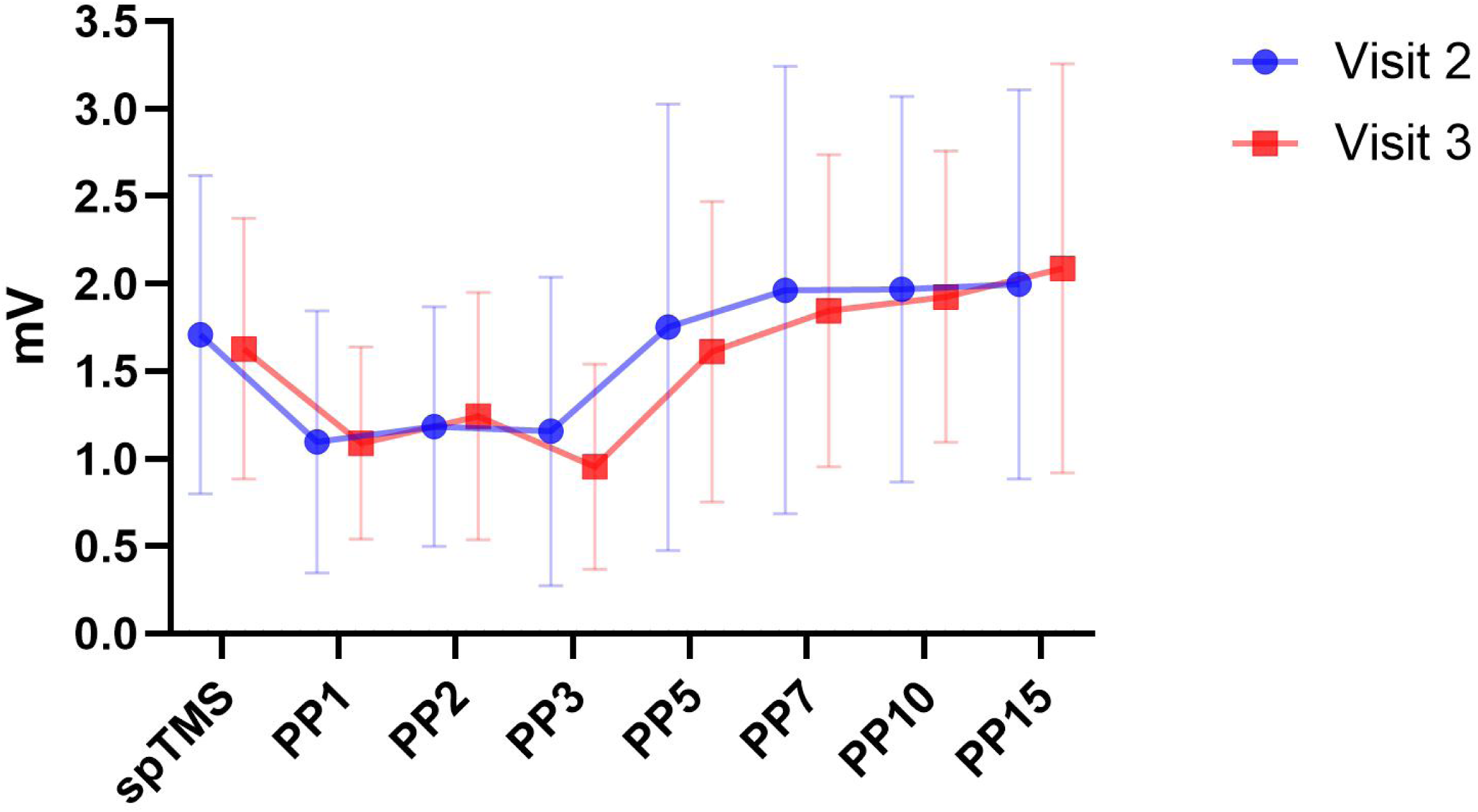
Test-Retest Reliability for the CIPN Group. *Mean with 95% Confidence Interval* *mV= millivolts; spTMS= single pulse at 130% of resting motor threshold; PP=paired pulse followed by the interstimulus interval value* Figure 4B. Test-Retest Reliability for the DN Group *Mean with 95% Confidence Interval* *mV= millivolts; spTMS= single pulse at 130% of resting motor threshold; PP=paired pulse followed by the interstimulus interval value*

**Table 3.** Intraclass Correlation Coefficient for Test-retest Reliability

| Measure | CIPN | DN | Total Sample |
| --- | --- | --- | --- |
| RMT | 0.60 (-0.52-0.89) | 0.97 (0.87-0.99) | 0.87 (0.67-0.95) |
| SP | 0.79 (0.28-0.94) | 0.98 (0.91-1.00) | 0.84 (0.58-0.94) |
| SICI | 0.84 (0.40-0.96) | 0.97 (0.85-0.99) | 0.87 (0.65-0.95) |
| ICF | 0.67 (-0.34-0.91) | 0.93 (0.65-0.99) | 0.73 (0.28-0.90) |
| *ICC values (95% confidence interval); CIPN= chemotherapy induced peripheral neuropathy; DN= diabetic neuropathy; RMT= resting motor threshold; SP= single pulse at 130% of resting motor threshold; SICI= short intracortical inhibition; ICF= intracortical facilitation* |  |  |  |

### Pre-Post change in TMS measures with active taVNS

All neuropathic pain symptoms, except for “shooting/electric shocks,” decreased post active taVNS (p< 0.05), and the greatest change was observed in tingling, numbness, and pain with median change values of -2.0, -2.0, and -1.5, respectively. It appears that there was a slight shift in the balance between SICI and ICF post active taVNS. SICI remained relatively unchanged from pre to post taVNS, with median values of -40.9% pre-taVNS (CI=-57.4 to -21.9) and -40.2% post-taVNS (CI=-61.3 to - 19.4). However, ICF decreased as indicated by median values of 10.1% pre-taVNS (CI=-4.8 to 36.4) and 1.5% post taVNS (CI=-6.3 to 19.0) (**Figure 5**).

**Figure 5.**
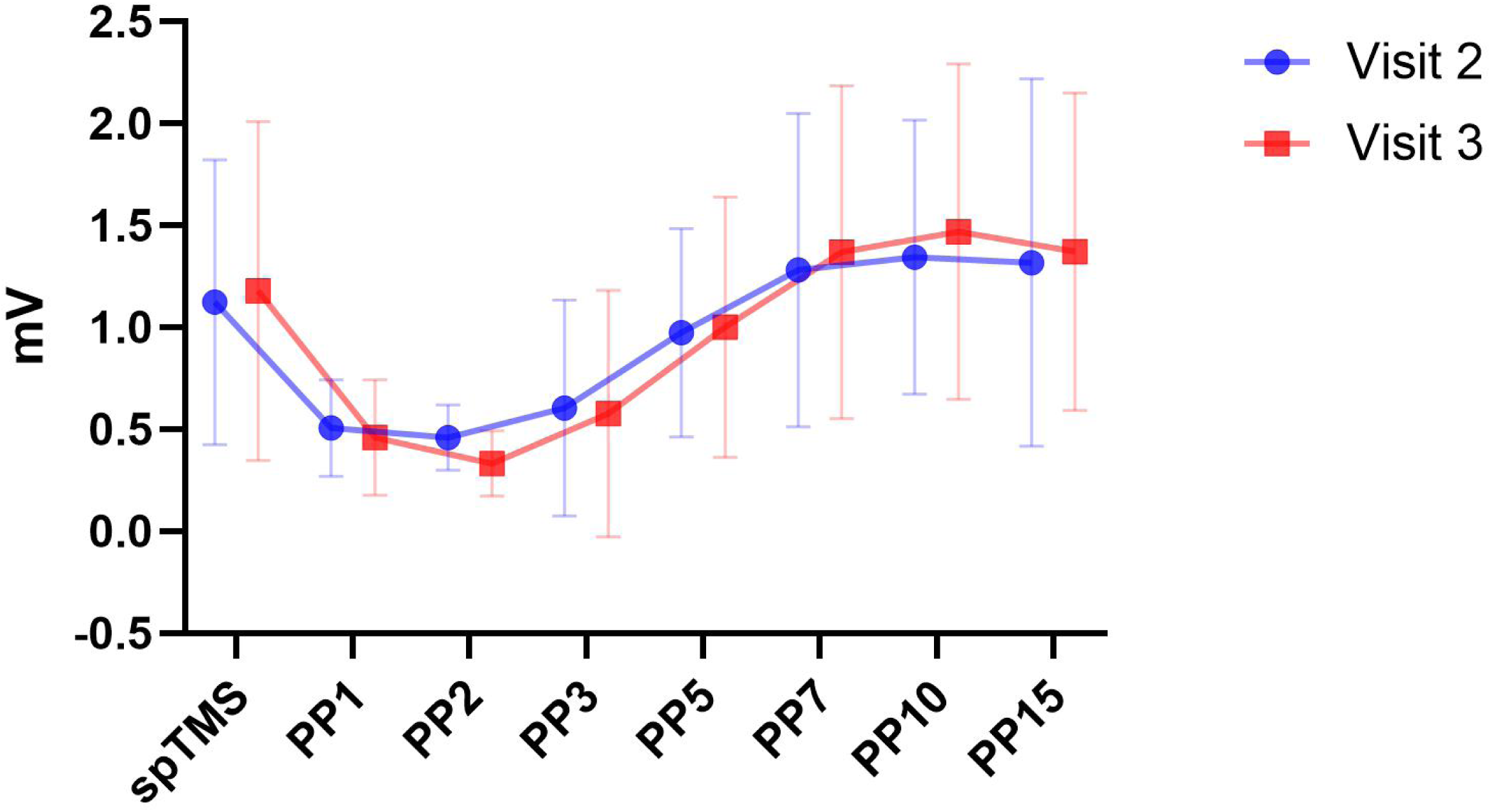
SICI and ICF Balance. *Median values with 95% Confidence Interval* *SICI=short intracortical inhibition; ICF=intracortical facilitation; spTMS=single pulse at 130% of resting motor threshold*

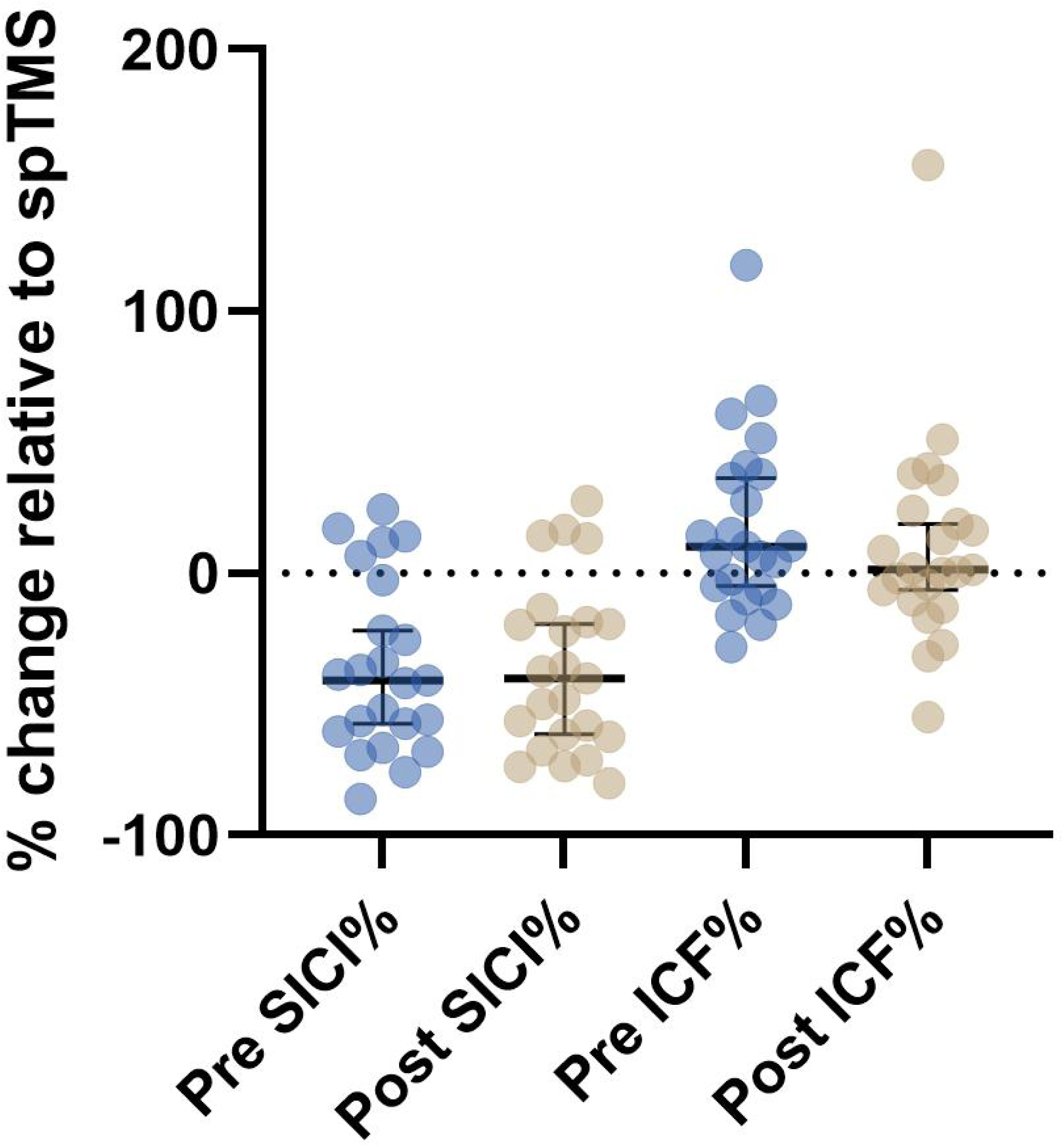

## DISCUSSION

In this secondary analysis study, we provide the first detailed description of TMS measures in people with painful PN associated with either neurotoxic chemotherapy exposure or diabetes. Importantly, the study sample consisted entirely of people who self-identified as Black or Hispanic/Latino, which are communities that are historically underrepresented in TMS research (32). Although hair type and style has been reported as a key barrier to conducting TMS research in Black communities (32), this was likely only a contributing factor for 2 of the 5 participants that we could not collect TMS data on. For these participants, the volume of hair prevented close contact of the coil with the scalp, and it is known that RMT increases linearly with the coil-cortex distance (47).

Thus, the high volume of hair between the coil and scalp likely elevated the required stimulus to achieve RMT beyond 77% of the maximum machine output, which was the ceiling for being able to complete the paired pulse protocol. However, 3 of the 5 participants with missing data had no hair or low hair volume, and we were still unable to achieve RMT within 77% of the maximum machine output.

### Comparison of our findings to previous research

Heterogeneity in TMS protocols across studies limit our ability to compare the findings from this study. However, RMT and SICI have been established as a stable measures with good reliability in healthy people over the age of 50 (48, 49) and in people with diabetes (50), and we found the same to be true in people with PN. One study found median values of -50% for SICI and 40% for ICF in healthy adults over 50 (49), which suggests that the sample of people with PN used for this study may have diminished capacity for both corticospinal inhibition and facilitation.

### Test-Retest Reliability

When assessing the entire sample, the test-retest reliability was good for all TMS measures. However, when separated by diagnoses, the ICC values were excellent for those with DN (≥0.93) and moderate-to-good for those with CIPN (0.60-0.84). Importantly, SICI had good reliability for both groups. The International Federation of Clinical Neurophysiology recently published a comprehensive update on the clinical diagnostic utility of TMS (23), and they concluded that of the TMS techniques, a reduction in SICI was most consistently associated with chronic pain, and therefore SICI might be used as a biomarker to select candidates for analgesic cortical neuromodulation (23). Our findings support the potential for using SICI as a biomarker.

### Response to taVNS

Significant changes in TMS measures were not expected in response to a single 60-minute session of taVNS, nor was the study adequately powered to assess such changes. Nevertheless, the description provided of SICI and ICF pre and post taVNS provides valuable preliminary data for the development of hypotheses on the mechanisms through which taVNS may impact PN as well as for the planning of future studies. Symptoms improved post active taVNS, and SICI did not change. However, there was a slight decrease in ICF, and this may suggest that taVNS induced a change in the balance between corticospinal inhibition and facilitation, towards inhibition. However, this is speculative and future research is warranted to investigate the relationships between SICI, ICF, and PN symptoms.

### Limitations

The findings of this study should be viewed as preliminary and interpreted with caution given the small sample size. Despite the limited generalizability, the fact that our entire sample consisted of Black and Hispanic/Latino people with PN adds value to the findings, as these communities have thus far been underrepresented in TMS research (32). Future studies are needed with diverse samples to determine the psychometric properties of these measures of people with PN.

### Conclusions

In this sample of Black and Hispanic/Latino people with PN, TMS measures were found to have good test-retest reliability. Additionally, we provided descriptive data on TMS measures that can be used for planning future studies to conclusively determine the psychometric properties and diagnostic utility of TMS measures for PN. This study establishes the feasibility of conducting TMS measures in this population, and the findings suggest that TMS measures, SICI in particular, may be promising tools for examining neurophysiological changes associated with PN and its treatment.

## Ethics statements

### Studies involving animal subjects

Generated Statement: No animal studies are presented in this manuscript.

### Studies involving human subjects

Generated Statement: The studies involving humans were approved by University of Miami Institutional Review Board. The studies were conducted in accordance with the local legislation and institutional requirements. The participants provided their written informed consent to participate in this study.

### Inclusion of identifiable human data

Generated Statement: No potentially identifiable images or data are presented in this study.

### Data availability statement

Generated Statement: The raw data supporting the conclusions of this article will be made available by the authors, without undue reservation.

### Generative AI disclosure

No Generative AI was used in the preparation of this manuscript.

### Scope Statement

This manuscript advances human neuroscience and fits with the scope of Frontiers in Human Neuroscience by providing novel data on transcranial magnetic stimulation (TMS) measures in people with peripheral neuropathy. Moreover, it provides data on TMS measures pre and post transcutaneous auricular vagus nerve stimulation, making it particularly well suited for the Research Topic “Non-invasive brain stimulation for chronic pain management.”

### Credit Author Statement

**Danylo F Cabral**: Methodology, Writing – original draft, Writing – review & editing. **Gabriel Gonzalez**: Investigation, Writing – original draft, Writing – review & editing. **Jessica Bolanos**: Investigation, Writing – original draft, Writing – review & editing. **Juan Gonzalez**: Investigation, Writing – original draft, Writing – review & editing. **Lisa McTeague**: Conceptualization, Funding acquisition, Methodology, Writing – original draft, Writing – review & editing. **Marlon L Wong**: Conceptualization, Data curation, Formal Analysis, Funding acquisition, Investigation, Methodology, Project administration, Resources, Software, Supervision, Validation, Visualization, Writing – original draft, Writing – review & editing. **Peter J Fried**: Conceptualization, Methodology, Writing – original draft, Writing – review & editing. **Peter J Hosein**: Funding acquisition, Investigation, Writing – original draft, Writing – review & editing.

## Data Availability

All data produced in the present study are available upon reasonable request to the authors

